# Saliva cell-free mitochondrial DNA (cf-mtDNA) as a dynamic biomarker of stress and emotion in daily life: Evidence from two independent repeated-measures studies

**DOI:** 10.64898/2026.03.23.26348537

**Authors:** Lauren Petri, Sun Ah Lee, David Shire, Samantha Leonard, Alexander Behnke, Jody Greaney, Lacy Alexander, David M. Almeida, Martin Picard, Caroline Trumpff

## Abstract

The present study analyzes the impact of naturalistic stress and emotions on saliva cell-free mitochondrial DNA (cf-mtDNA) in daily life across two independent cohorts with different temporal resolutions. Study 1 examined the interaction between daily stress and major depressive disorder (MDD) on cf-mtDNA in young adults (n= 18, 8 MDD, 10 controls) across four days. For individuals with MDD, stress exposure was associated with a 68% reduction in cf-mtDNA. A higher number or greater severity of stressors also reduced cf-mtDNA by 24 to 27%. Study 2 extended this framework by implementing a finer temporal resolution, measuring saliva and affective states every hour, up to 20 times per day for 2 days (n = 25). Negative emotions, including stress and frustration, were associated with reductions in cf-mtDNA of 15%, whereas positive emotions, such as happiness and calm, predicted increases of up to 28%. The strength and direction of the effects were person- and context-dependent. These findings suggest that cf-mtDNA does not exhibit a uniform stress response in daily life. Instead, it reflects dynamic signaling shaped by timing, emotional context, and diagnostic status. This work demonstrates that interpreting cf-mtDNA as a stress biomarker in real-world settings requires modeling timing and heterogeneity of effects.

**Highlights:**

- Saliva cf-mtDNA dynamically tracks with psychosocial experience in real-world contexts.
- Daily stress was associated with marked cf-mtDNA reductions in MDD, revealing stress-contingent vulnerability.
- Higher stressor load was associated with dose-dependent suppression of cf-mtDNA.
- At the hourly level, negative emotions predicted lower cf-mtDNA, whereas positive emotions predicted higher levels.
- cf-mtDNA showed greater sensitivity to psychosocial experience than cf-nDNA, supporting the notion that mitochondrial DNA release is regulated beyond passive cell death.

## Introduction

Stress is among the most potent predictors of poor mental and physical health outcomes, conferring an increased risk of depression, obesity, diabetes, and cancer (Epel et al., 2018). In everyday life, stress can arise from relatively minor yet frequent events, such as tight work deadlines and interpersonal conflicts, which occur on approximately 40% of days in community samples (Almeida et al., 2002). Although it is well established that repeated activation of the stress response is a major contributor to accelerated physiological wear and tear (McEwen, 1998), the rapid biological cascades that follow naturally occurring stressors have not been fully characterized. Advances in stress assessment methods, particularly ecological momentary assessment (EMA) and daily diary approaches, have enabled detailed characterization of these stress responses as they unfold in real time (Almeida et al., 2002; Shiffman et al., 2008). However, measuring relevant biomarkers at comparable frequencies has proven difficult, severely limiting our understanding of the immediate biological effects of naturalistic stressors. Addressing this gap will require the use of biomarkers that can be sampled frequently in daily life, aligning with the temporal granularity of existing psychological assessments.

### 1.1 Cell-free mitochondrial DNA as a candidate biomarker of acute stress processes in daily life

Cell-free mitochondrial DNA (cf-mtDNA) is a compelling candidate to address this gap. While cortisol is one of the most commonly studied stress biomarkers (Hellhammer et al., 2009), it only reflects the activity of a single arm (i.e., the hypothalamic-pituitary-adrenal axis) of a much broader stress-response network (Picard & McEwen, 2018). As central regulators of cellular energy and adaptation, mitochondria represent another essential arm, or potential downstream target of this system (Picard & McEwen, 2018). Following stress, mitochondria release mitochondrial-derived molecules, like cf-mtDNA, into circulation, which has systemic implications. MtDNA shares characteristics (e.g., circularity and size) with bacterial DNA that can trigger an inflammatory response when released into circulation (Collins et al., 2004; West & Shadel, 2017).

This immunological activity has positioned cf-mtDNA as a pro-inflammatory biomarker of disease (de Miranda et al., 2024; Harrington et al., 2019; Meng et al., 2019; Sayal et al., 2023; Kananen et al., 2020). Elevated levels are consistently associated with poor health outcomes in severe illnesses like cancer and sepsis (de Miranda et al., 2024; Harrington et al., 2019; Meng et al., 2019; Sayal et al., 2023; Kananen et al., 2020). However, emerging evidence suggests cf-mtDNA is not merely a marker of acute pathology. It fluctuates dynamically, exhibiting diurnal variation and rapid reactivity to psychosocial stress (Limberg et al., 2025; Trumpff et al., 2019, 2022, 2025; Behnke et al., 2025). In controlled experiments, serum cf-mtDNA increases ∼1.7 to 3-fold within 20-30 minutes of socio-evaluative stress (Hummel et al., 2018), and saliva cf-mtDNA levels rise by ∼2.8-fold on average (with some participants showing >10-fold elevations) within just 10 minutes (Trumpff et al., 2025). These findings suggest that cf-mtDNA may function as a rapid stress-responsive signal, rather than solely as a downstream correlate of disease. Determining whether cf-mtDNA exhibits comparable responsiveness to real-world stressors is critical for clarifying its role in the biological embedding of stress (Trumpff et al., 2019).

### 1.2 Psychiatric conditions and cf-mtDNA

Chronic stress disrupts saliva cf-mtDNA dynamics, predicting flattened cf-mtDNA rhythms and reduced morning surges 1-4 hours after awakening (Behnke et al., 2025). Altered stress responsivity is a hallmark of many psychopathologies, and decades of research on cortisol show that both reactivity and recovery differ systematically among these individuals (Burke et al., 2005; Zorn et al., 2017). Among psychiatric disorders, major depressive disorder (MDD) stands out as a particularly relevant context in which to study potential alterations to cf-mtDNA-stress dynamics (Burke et al., 2005; Zorn et al., 2017). MDD is characterized by persistent low mood, altered reactivity to daily stress, impairments in energy metabolism, and increased oxidative stress (Gimenez-Palomo et al., 2021). Accordingly, individuals with MDD may exhibit distinctive cf-mtDNA release patterns, particularly following acute stress. Studies comparing cf-mtDNA concentrations between groups have yielded mixed results, underscoring the need for within-person designs (Lindqvist et al., 2016, 2018; Melamud et al., 2023; Park et al., 2022; Behnke et al., 2023; Trumpff et al., 2021).

### 1.3 The present study

In the present study, we examine whether saliva cf-mtDNA responds to day- and momentary-level psychological stress under ecologically valid, real-world conditions, using data from two independent cohorts with intensive repeated-measures designs. Across both studies, we test whether real-world psychosocial dynamics map onto cf-mtDNA in the same direction as laboratory stress reactivity, or whether timing and psychological context produce distinct signatures. Study 1 investigates whether exposure to daily stress influences saliva cf-mtDNA concentrations among healthy adults and individuals with MDD across four consecutive days. Study 2 uses hourly sampling with EMA to examine how acute fluctuations in stress and emotion affect concurrent saliva cf-mtDNA concentrations. The primary outcome of the study is cf-mtDNA; however, cell-free nuclear DNA (cf-nDNA) is also quantified as a control.

Divergence of these markers would suggest a more specific regulatory mechanism outside of cell death. Additionally, while stress is the primary focus of this paper, affect represents a broader, multidimensional construct that both shapes and is shaped by stress. Negative affect can arise as a consequence of stress, yet heightened negative affective states may also amplify the perception of stress (Feldman et al., 1999). Because positive affect can influence physiological processes and may buffer stress responses (van Steenbergen et al., 2021), we conducted exploratory analyses testing whether cf-mtDNA is sensitive not only to stress and negative affect individually, but also to their interplay with positive affect.

## 2. Methods

### 2.1. Study 1: Daily Stress, cf-mtDNA, and major depressive disorder

#### 2.1.1 Participants and procedures

This study was approved by The Pennsylvania State University (IRB #9617) and registered with the Food and Drug Administration (IND #125994). The full protocol has been previously described in detail (Greaney et al., 2020; study design in Figure 1A). Participants (n = 18, 8 MDD, 10 healthy controls [HC]; 10 females; age: *M* = 23 ± 4 years; BMI: *M* = 24.95, *SD* = 4.76 kg/m^2^) underwent the Mini-International Neuropsychiatric Interview (MINI), which identifies current or past major psychiatric disorders according to the criteria in the Diagnostic and Statistical Manual of Mental Disorders (DSM–5). The MINI diagnostic algorithm identified 8 participants who met criteria for MDD (4 female) and 10 healthy controls (6 female) who demonstrated no history of major psychiatric illness (Sheehan et al., 1998). Exclusion criteria included the presence of any comorbid major psychiatric condition (e.g., bipolar disorder, schizophrenia, etc.), the current or recent (within 8 weeks) use of psychoactive or psychopharmacological drugs (including antidepressants and anxiolytics), and active suicidal ideation with intent or plans. Additional details regarding diagnostic classification and clinical screening procedures can be found in previous publications (Darling et al., 2024; Greaney et al., 2019, 2022).

**Figure 1.**
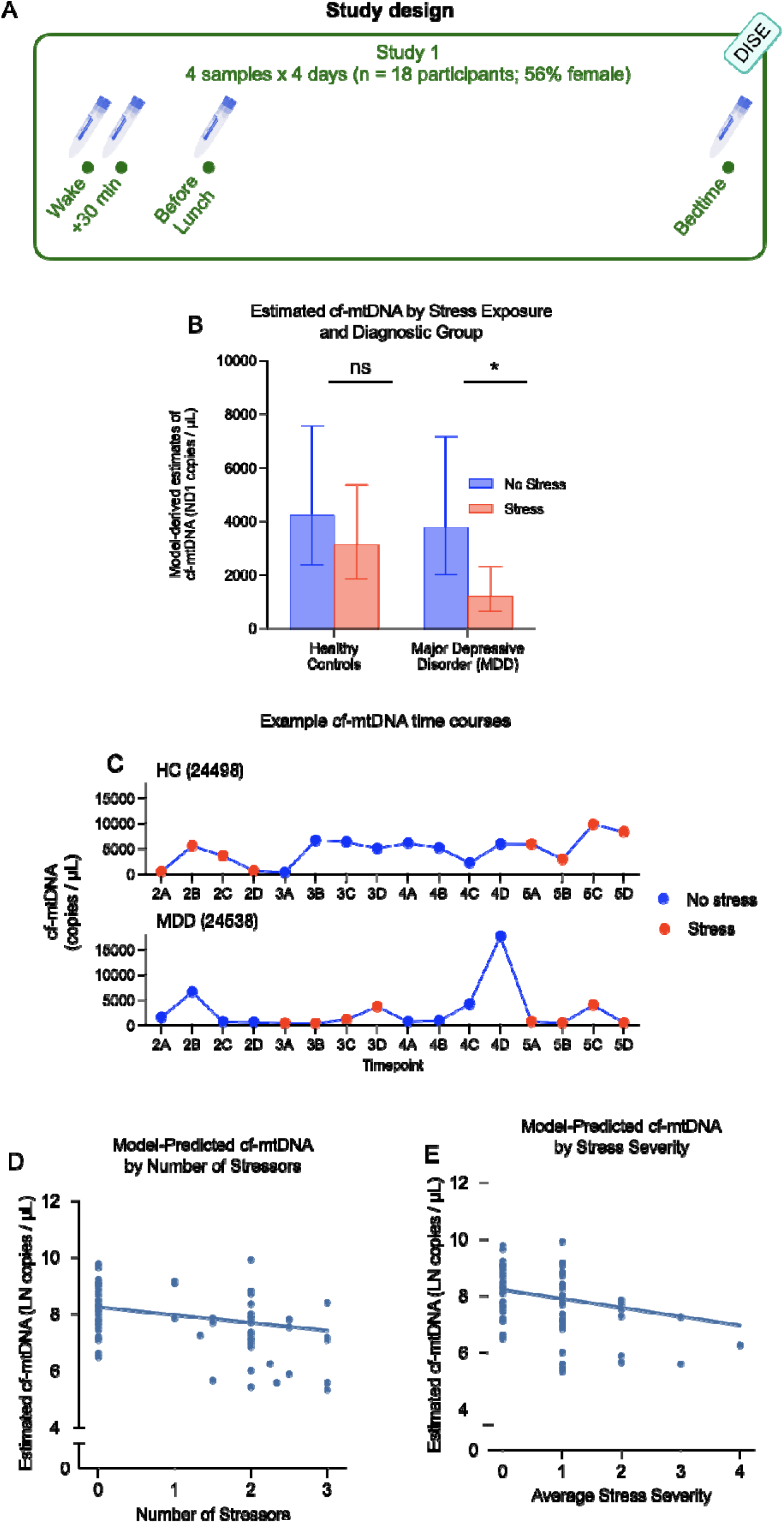
Study 1: Cell-free mitochondrial DNA (cf-mtDNA) dynamics by day-level stress. **(A)** Protocol schematic depicting four saliva samples per day across four consecutive days with nightly Daily Inventory of Stressful Events (DISE) interviews (n = 18, MDD = 8, HC = 10, 260 saliva samples). **(B)** The interaction between cf-mtDNA and stress exposure for healthy controls and participants with Major Depressive Disorder (MDD). The effect of stress exposure was significant for participants with MDD (68% reduction), but not for healthy controls. **(C)** Example plot of cf-mtDNA concentrations across all timepoints and days for one participant with MDD and one healthy control. On the x-axis, the numeric value represents the day, and the letter represents the time point within the day. (**D)** Model-predicted values of cf-mtDNA as a function of the number of stressors corresponding to a 27% cf-mtDNA decrease (log-*b* = -0.31*, p* = .01)**. (E)** Model-predicted means with 95% CI demonstrating each one-unit increase in stress severity is associated with a 24% lower cf-mtDNA level.

#### 2.1.2 Saliva collection

Participants collected saliva four times each day (i.e., immediately upon awakening, 30 minutes after awakening, before lunch, and at bedtime) for four consecutive days. Participants were provided with detailed instructions to collect samples independently at home using Sarstedt Salivettes (Cat# 51.1534.500). Participants recorded saliva collection times in daily diaries. Samples were stored in participants’ home freezers at approximately –20°C. The samples were transported on ice to the Mitochondrial Psychobiology Laboratory at Columbia University for processing. Nearly all participants returned complete sample sets (16 samples each), totaling 284 samples or 72 complete days of data. Six days (24 samples) were excluded due to missing stress data, and one additional day (4 samples) was removed for being outside the interquartile range for cf-mtDNA. After exclusions, 260 samples or 65 days of data (29 non-stressor and 36 stressor) were included in the final analysis.

#### 2.1.3 Assessment of daily stress

Daily stress was measured using the Daily Inventory of Stressful Events (DISE), a validated 24-hour retrospective checklist of six common stressors: arguments, argument avoidance, stressful events at work or school, stressful events at home, discrimination, network stress, and any other stressors (Almeida et al., 2002). A day was classified as a stressor day if at least one DISE-validated event was reported, and as a non-stressor day if no events occurred. Participants rated the perceived severity of each stressor on a 0–4 scale, which was averaged to yield a daily mean severity score. Additionally, the total number of stressors reported was summed to capture the frequency of daily stressors. Saliva concentrations were averaged within day to align biomarker timescales with the DISE, yielding day-level cf-mtDNA and cf-nDNA values.

### 2.2. Study 2: Hourly affect and cf-mtDNA

#### 2.2.1 Participants and procedures

This study was approved by the New York State Psychiatric Institute Institutional Review Board (IRB #8149). Adults (n = 25, 13 females, age: *M* = 31 ± 8 years; BMI: *M* = 23.9*, SD* = 3.1 kg/m^2^) were recruited from the Columbia University Irving Medical Center (CUIMC) regional area (Behnke et al., 2025). Participants completed two days of intensive hourly saliva and EMA sampling according to established procedures (Kirschbaum & Hellhammer, 1994; see study design in Figure 2A).

**Figure 2.**
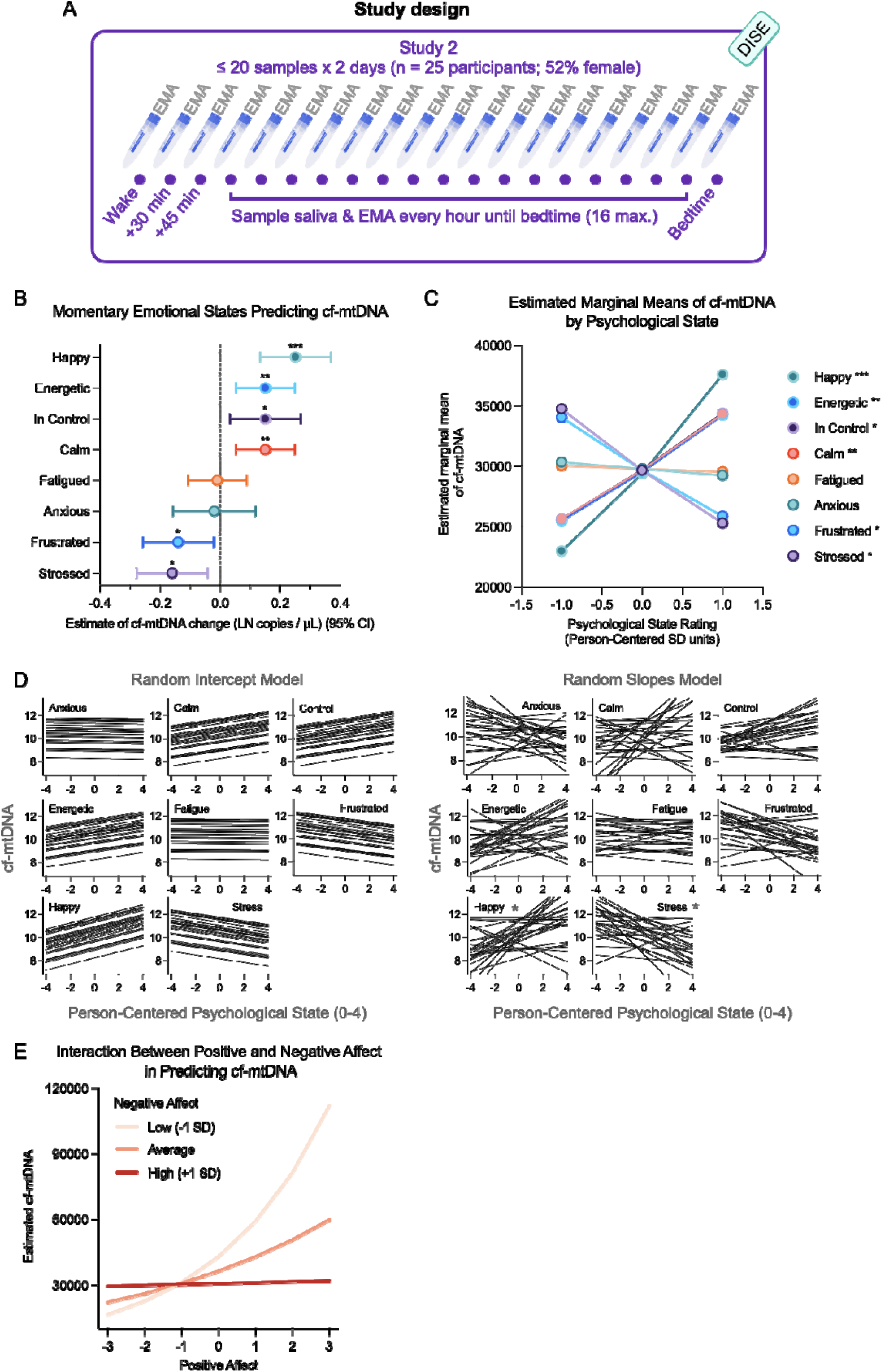
Study 2: Cell-free mitochondrial DNA (cf-mtDNA) dynamics by momentary emotional states. **(A)** Protocol schematic depicting two days of hourly saliva collection with simultaneous EMAs (n = 25, 826 saliva samples). **(B)** Forest plot of fixed effects from gamma generalized linear mixed models (random intercepts; each emotion was person-centered). Asterisks indicate significant effects. **(C)** Estimated marginal means of cf-mtDNA levels on the response scale (copies/μL) plotted by psychological state (+/- 1 SD), error bars = 95% CI. **(D)** Models with fixed and random slopes demonstrate the person-specific heterogeneity of effects, with asterisks indicating significant effects for happiness (-49% to + 262%) and stress (-68% to 80%). Only 6 participants had negative slopes for happiness, while 4 participants had positive slopes for stress. **(E)** Positive Affect (PA) x Negative Affect (NA) interaction using CFA-derived latent factors. At the mean level of NA, higher PA was associated with higher cf-mtDNA (log-*b* = .30, SE = 0.11, *p* = .008), whereas NA showed no main effect at the mean level of PA (log-*b* = –0.04, SE = 0.09, *p* = .63). No significant main effects or interaction were observed for cf-nDNA (log-*b* = – 0.15, SE = 0.11, *p* = .16). Thus, higher NA progressively attenuated the positive association between PA and cf-mtDNA.

#### 2.2.2 Saliva collection

On each study day, participants collected three saliva samples within the first hour of awakening (immediately upon awakening, 30 minutes post-awakening, and 45 minutes post-awakening), followed by hourly samples for the remainder of the day, yielding up to 20 samples per day. To minimize contamination, participants were instructed to refrain from eating, brushing their teeth, and drinking for at least 30 minutes before collection. Samples were collected using Sarstedt Salivettes (Cat# 51.1534.500), stored in home freezers at approximately –20°C and shipped on ice to the Mitochondrial Psychobiology Laboratory at Columbia University for cf-mtDNA analysis. On average, participants returned 16.5 (*SD* = 3.68) saliva samples per day. A single outlier with a cf-mtDNA value 16 SDs above the mean was removed; however, analyses were similar without it (see Supplemental Table S1).

#### 2.2.3 Assessment of emotional states

During each saliva collection, participants completed a brief survey on a mobile device through REDCap, rating their current emotional state. The EMA included ratings of eight emotions selected to capture both positive (i.e., happy, energetic, in control, and calm) and negative (i.e., stressed, anxious, fatigued, and frustrated) affective states. Responses were recorded on a 5-point scale (0 = “not at all” to 4 = “extremely”). On average, participants completed 13.86 (*SD* = 3.17) EMA measurements per day. Ratings were person-centered, so coefficients reflected changes in cf-mtDNA for a 1-unit deviation from each individual’s average. Person-centered emotion ratings were analyzed separately in bivariate models. Non-person-centered values were used in a confirmatory factor analysis (CFA) to derive composite factor scores for momentary positive affect (PA) and negative affect (NA). These scores were used to test the effect of PA, NA, and their interaction on cf-mtDNA.

#### 2.3.1 Saliva processing for Study 1 and Study 2

For both studies, saliva samples were stored at –80 °C and processed in batches to minimize inter-assay variability. Salivettes were thawed at room temperature and centrifuged at 1,000 g for five minutes. The clear saliva fraction was carefully aspirated from the top of the tube using a pipette, leaving the cellular debris at the bottom of the tube. This fraction was transferred to 1.5 mL microcentrifuge tubes and spun again at 5,000 g for 10 minutes. The resulting supernatants were transferred to new 1.5 mL tubes.

#### 2.3.2 Cf-mtDNA and cf-nDNA quantification

Cf-mtDNA and cf-nDNA were quantified from the saliva supernatant using a modified version of the MitoQuicLy protocol (Michelson et al., 2023). Supernatants were thermolyzed in duplicate overnight. The resulting lysates were analyzed in triplicate using real-time quantitative polymerase chain reactions (qPCR) with TaqMan chemistry, targeting the mitochondrial gene ND1 and the nuclear gene B2M simultaneously. For each target, triplicate cycle threshold (Ct) values were compared against a DNA standard curve to calculate absolute copy numbers. Triplicate Ct values were inspected, and the two most concordant replicates were averaged to obtain the final value. Between-plate reliability of duplicates was tested using intraclass correlation coefficients (ICCs) with the irr package in R (Gamer et al., 2019; version 4.4.2; R Core Team, 2024).

### 2.4. Analytical approach

#### 2.4.1 Study 1

Prior to conducting the primary analysis, we ensured the number and severity of stressors did not differ by diagnostic group (MDD vs. HC). To test this, we used a Poisson generalized linear mixed-effects model with a log link for the number and a linear mixed-effects model for severity. Additionally, we assessed the relationship between log-transformed cf-mtDNA and cf-nDNA concentrations using repeated measures correlations. To examine the influence of stressor days, diagnostic group, and their interaction on saliva cf-mtDNA levels, generalized linear mixed-effects models (GLMMs) with a gamma error distribution, log link, and random intercepts to capture individual differences in baseline cf-mtDNA were computed using the glmmTMB package in R (Brooks et al., 2017). The primary model was specified as:

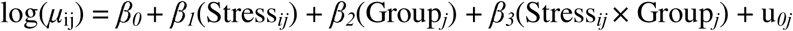

where *μ*_ij_ represents the expected cf-mtDNA concentration for participant *j* on day *i*, *β_0_* is the fixed intercept, and u0j *N*(0, σ^2^_u_) captures the between-person variability in baseline cf-mtDNA levels (no stress day, number of stressors = 0, or stress severity of 0). *β_2_* estimates the differences in cf-mtDNA levels between diagnostic groups, while *β_3_* captures the interaction effect, indicating whether stress-related changes in cf-mtDNA differ between MDD and HC participants. The second model incorporated time-invariant covariates (age, sex, and BMI) to control for potential confounders:

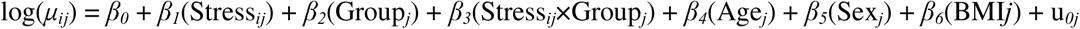

where *β_4_*, *β_5_*, and *β_6_* estimate the effects of age, sex, and BMI, respectively. Additionally, both models were performed with cf-nDNA as the outcome. These models served as an additional check to determine whether the observed cf-mtDNA relationship was specific to mtDNA or reflected broader changes (e.g., cell death). A significant effect of stress on cf-mtDNA, but not cf-nDNA, would suggest mitochondrial specificity.

Model fit was assessed using Akaike Information Criterion and Bayesian Information Criterion (AIC/BIC). Lower relative AIC and BIC values indicate a better fit, and a reduction of greater than 2 points was interpreted as a meaningful improvement (Burnham & Anderson, 2004). Estimated marginal means and contrast estimates were calculated with the emmeans package to obtain back-transformed effects on the original response scale (Lenth, 2025).

#### 2.4.2 Study 2

Study 2 extended the analytical framework of Study 1 by modeling fluctuations in eight momentary person-centered psychological states as predictors of corresponding cf-mtDNA levels. In contrast to Study 1, which utilized day-level averages and 24-hour retrospective daily diaries, this study used the individual momentary assessments and cf-mtDNA and cf-nDNA levels. The effects of each of the EMA psychological variables on cf-mtDNA and cf-nDNA were analyzed using GLMMs with gamma error distributions, log links, and random intercepts to capture individual differences in baseline cf-DNA. The primary model assessed the relationship of person-centered psychological states and cf-mtDNA while allowing for between-person variability in baseline cf-mtDNA through random intercepts:

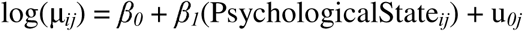

We conducted a CFA to reduce the eight psychological states into two latent affective dimensions: Negative Affect (NA), which included the variables stressed, anxious, fatigued, and frustrated, and Positive Affect (PA) which included the variables calm, in control, energetic, and happy. The model was estimated using full-information maximum likelihood (FIML). Modification indices were examined to improve model fit, and changes were implemented iteratively. Residual covariances between energetic and happy, as well as between fatigued and energetic, were added based on these indices. Additionally, a cross-loading of fatigue onto both PA and NA was introduced. All modifications were made one at a time and retained only if they improved model fit without undermining theoretical interpretability. The final model demonstrated a good fit (*χ^2^* (16) = 92.48, *p* < .001; CFI = 0.98; RMSEA = 0.07; SRMR = 0.03). All factor loadings were statistically significant (*ps* ≤ .006). Loadings ranged from 0.13 to 0.69 for NA and from 0.85 to 1.21 for PA, with fatigued cross-loading negatively onto PA. Factor scores derived from the CFA were grand-mean centered. Those values were entered into a GLMM as predictors to test whether associations with cf-mtDNA were dependent on concurrent emotions while accounting for between-person variability via random intercepts using the following equation:

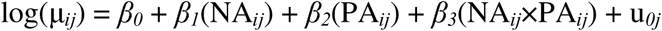

## 3. Results

### 3.1. Study 1: Daily Stress, cf-mtDNA, and major depressive disorder

#### 3.1.1 Sample characteristics and data quality

On stressor days, participants reported approximately one stressor (*M* = 1.39, *SD* = 0.73, range 0–4) with an average severity of 1.89 (*SD* = 0.78; range 0–3). Notably, participants with MDD did not differ from healthy controls in the number (log-*b* = 1.13, 95% CI [0.50, 2.31], *p* = .73) or severity (log-*b* = –0.08, 95% CI [–0.83, 0.65], *p =* .83) of stressors reported.

Between-plate reliability was excellent for cf-mtDNA (ICC = .95, *p* < .001) and cf-nDNA (ICC = .93, *p* < .001). Within persons, cf-mtDNA and cf-nDNA were positively correlated (*r_rm_ =* .59, 95% CI [0.49, 0.67], *p* < .001; Supplemental Figure S1A), reflecting the expected moderate concordance between the two markers.

#### 3.1.2 Stress exposure is associated with reduced cf-mtDNA

Daily stress exposure predicted lower cf-mtDNA in MDD, suggesting stress-contingent dysregulation rather than a baseline group difference (log-*b* = –0.85, 95% CI [–1.65, –0.05], *p* = .03; Table 1 Model 1; Figure 1B). Post hoc comparisons showed that in healthy controls, cf-mtDNA levels were approximately 26% lower on stressor days (∼3,145 copies/µL) versus non-stressor days (∼4,238 copies/µL), although this difference was not significant (1.35-fold; log-contrast = 0.30, SE = 0.25, *p =* .22). In individuals with MDD, cf-mtDNA concentrations were 68% lower on stressor days (∼1,206 copies/µL) versus non-stressor days (∼3,791 copies/µL; log-contrast = 1.15, SE = 0.32, *p* < .001). Example trajectories for MDD and HC participants are shown in Figure 1C.

**Table 1.**
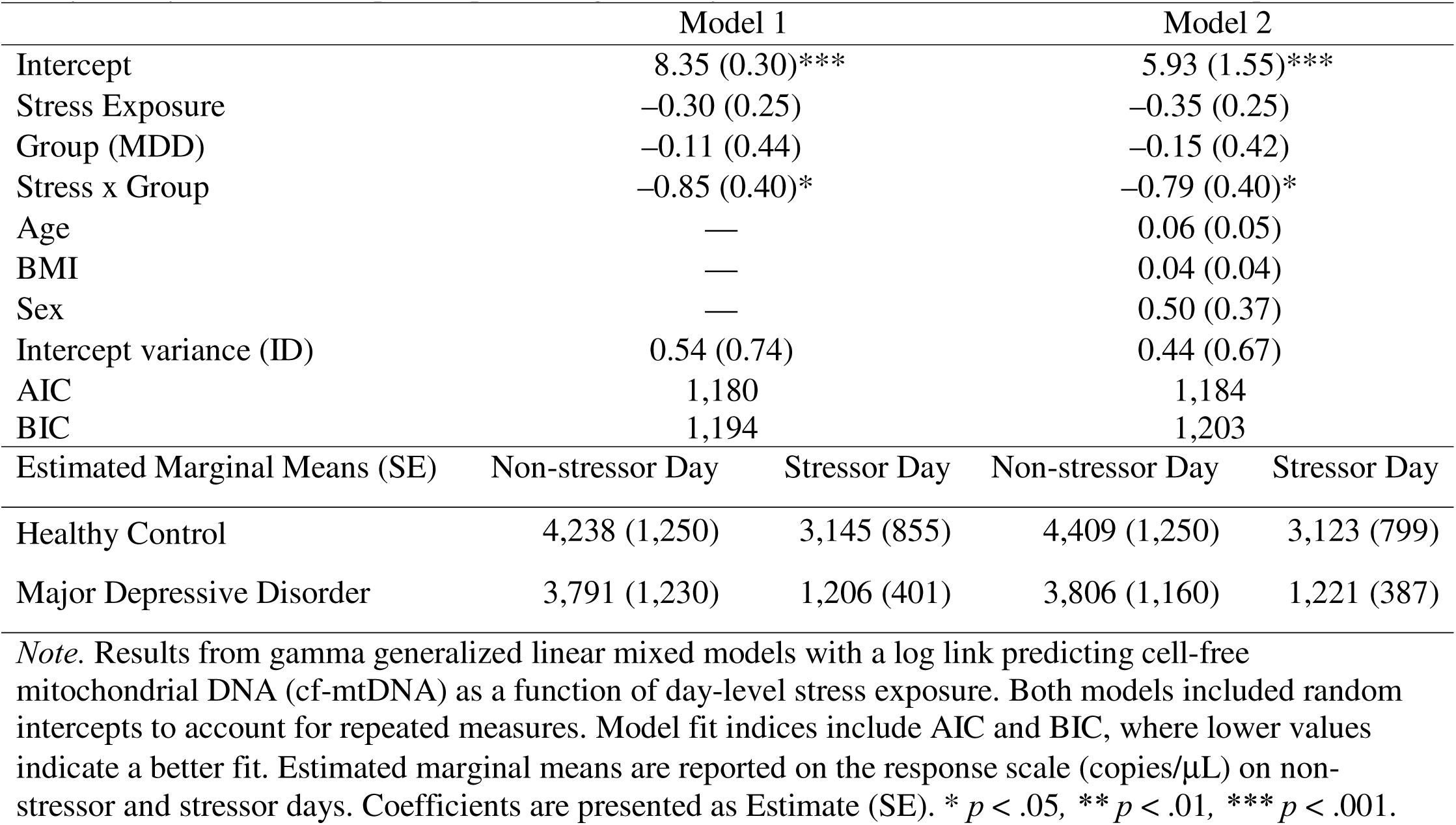
Study 1: Day-level stress exposure predicting saliva cf-mtDNA in GLMMs with random intercepts.

The conditional R^2^ suggests this model explained approximately 71% of the variance, with the marginal R^2^ (fixed effects) accounting for 20.4%, indicating substantial inter-individual variability in cf-mtDNA. A sensitivity analysis adjusting for BMI, age, and sex did not change the pattern of results (Table 1, Model 2). These covariates collectively added ∼10% more explained variance through fixed effects, but none were individually significant. Although cf-nDNA correlated with cf-mtDNA within persons, there was no evidence of a relationship between stress exposure and cf-nDNA (*p*s > .25; Supplemental Figures S2A–S2C, Supplemental Table S2), suggesting that the effect of stress was specific to cf-mtDNA.

#### 3.1.3 A higher number and severity of stressors were associated with lower cf-mtDNA

An exploratory analysis of the dose-response effect of stress showed that the number (Figure 1D) and severity (Figure 1E; Supplemental Table S3) of stressors were each associated with lower cf-mtDNA: Each additional stressor experienced was associated with a 27% reduction (log-*b* = –0.31, SE = 0.12, *p* = .01) in cf-mtDNA at the day level. Similarly, a one-unit increase in stressor severity was associated with a 24% decrease (log-*b* = –0.28, SE = 0.10, *p* = .004) in cf-mtDNA. Although these models demonstrate a dose-response effect of stress on cf-mtDNA, we consider these results preliminary due to the limited number of high-stress scores in this cohort.

### 3.2. Study 2: Hourly affective states and cf-mtDNA

#### 3.2.1 Sample characteristics and data quality

A total of 826 saliva samples were returned. Data for seven emotions (i.e., happy, energetic, calm, in control, anxious, fatigued, and frustrated) were available for 807 samples, while 762 samples had valid stress ratings. Among positive emotions, calm was reported most often (present on 88.7% of assessments, *M* = 2.24, *SD* = 1.25), followed by feeling in control (85.7%, *M* = 2.11, *SD* = 1.31), happy (81.2%, *M* = 1.75, *SD* = 1.15), and energetic (74.3%, *M* = 1.47, *SD* = 1.19). Negative emotions were reported less frequently, with fatigue present on 54.4% of assessments (*M* = 0.88, *SD* = 1.05), followed by stress (41.5%, *M* = 0.56, *SD* = 0.79), anxiety (36.0%, *M* = 0.49, *SD* = 0.75), and frustration (23.5%, *M* = 0.33, *SD* = 0.69).

Between-plate reliability was high for both cf-mtDNA (ICC = .99, *p <* .001) and cf-nDNA (ICC = .99, *p <* .001). Repeated measures correlation tests revealed that cf-mtDNA and cf-nDNA were strongly associated (*r_rm_* = .81, *p* < .001; Supplemental Figure S1B). At the hourly level, cf-mtDNA and cf-nDNA also fluctuated substantially.

#### 3.2.2 Cf-mtDNA increases with positive emotions and decreases with negative emotions

GLMMs with a gamma error distribution, a log link function, and random subject intercepts revealed divergent associations between person-centered affect ratings and contemporary cf-mtDNA concentrations (Table 2; Figure 2B–2C). Each of the four positive emotions was associated with higher cf-mtDNA levels. Every one unit increase in happiness corresponded to a 28% increase in momentary cf-mtDNA (log-*b* = 0.25, SE = 0.06, *p <* .001). Additional units in calmness (log-*b* = 0.14; SE = 0.05, *p* = .008), energy (log-*b* = 0.15, SE = 0.05, *p* = .002), and feeling in control (log-*b* = 0.15, SE = 0.06, *p* = .01) were each associated with ∼16% increases in saliva cf-mtDNA levels.

**Table 2.** Study 2: Person-centered psychological states predicting cf-mtDNA in GLMMs with random intercepts.

|  | Anxious | Calm | In Control | Energetic | Fatigued | Frustrated | Happy | Stressed |
| --- | --- | --- | --- | --- | --- | --- | --- | --- |
| Fixed Effects |  |  |  |  |  |  |  |  |
| Intercept | 10.30<br>(0.20)*** | 10.30<br>(0.20)*** | 10.30<br>(0.20)*** | 10.29<br>(0.20)*** | 10.30<br>(0.20)*** | 10.30<br>(0.20)*** | 10.29<br>(0.20)*** | 10.30<br>(0.21)** |
| Variable | -0.02<br>(0.07) | 0.14<br>(0.05)** | 0.15<br>(0.06)** | 0.15<br>(0.05)** | -0.01<br>(0.05) | -0.14<br>(0.06)* | 0.25<br>(0.06)*** | -0.16 (0.06) |
| Random Effects |  |  |  |  |  |  |  |  |
| Intercept | 1.00 (1.00) | 0.98<br>(0.99) | 0.98<br>(0.99) | 0.99<br>(0.99) | 1.00<br>(1.00) | 1.00<br>(1.00) | 0.97 (0.98) | 1.05 (1.00) |
| Model Fit Indices |  |  |  |  |  |  |  |  |
| AIC | 18,318 | 18,311 | 18,312 | 18,309 | 18,318 | 18,314 | 18,299 | 17,290 |
| BIC | 18,337 | 18,330 | 18,330 | 18,328 | 18,337 | 18,332 | 18,318 | 17,309 |
| Estimated Marginal Means (SE) |  |  |  |  |  |  |  |  |
| Low (-1) | 30,339 | 25,639 | 25,637 | 25,500 | 30,056 | 34,066 | 22,978 | 34,769 |
| Average<br>(0) | 29,796 | 29,666 | 29,702 | 29,562 | 29,797 | 29,685 | 29,400 | 29,660 |
| High (+1) | 29,263 | 34,327 | 34,410 | 34,272 | 29,540 | 25,868 | 37,616 | 25,301 |

*Note.* Results from gamma generalized linear mixed models with a log link predicting cell-free mitochondrial DNA (cf-mtDNA) as a function of person-centered emotions. All models included random intercepts to account for repeated measures. Model fit indices include AIC and BIC, where lower values indicate a better fit. Estimated marginal means are reported on the response scale (copies/μL) at low, average, and high levels of the psychological state. Coefficients are presented as Estimate (SE). \* $p < .05$ , \*\* $p < .01$ , \*\*\* $p < .001$ .

Conversely, higher stress and frustration ratings were linked to lower saliva cf-mtDNA concentrations. Every one unit increase in stress corresponded to a 15% decrease in cf-mtDNA (log-*b* = –0.16, SE = 0.06, *p =* .01), and every one unit increase in frustration corresponded to a 13% decrease in cf-mtDNA (log-*b* = –0.14, SE = 0.06, *p* = .03). Neither anxiety (*p* = .80) nor fatigue (*p* = .86) was significantly associated with momentary cf-mtDNA concentrations. Effects persisted after adjustment for age, sex, and BMI (Supplemental Table S4) and were robust to the inclusion of a statistical outlier (Supplemental Table S5).

#### 3.2.3 Person-specific heterogeneity in affect−cf-mtDNA coupling

Models including random slopes revealed between-person heterogeneity in the relationship of cf-mtDNA with happiness and stress perception (Figure 2D; Supplemental Table S6). In general, momentary cf-mtDNA levels were still positively associated with happiness ratings (log-*b* = 0.31, SE = 0.12, *p* = .009) and negatively associated with stress (log-*b* = –0.28, SE = 0.12, *p* = .02). However, the magnitude and, in some cases, the direction of within-person slopes varied widely across individuals (–49% to +262% for happiness, –68% to +80% for stress), again illustrating large potential inter-individual differences in these psychobiological associations.

#### 3.2.4 Interaction between positive and negative affect on cf-mtDNA

Confirmatory factor analysis supported a two-factor model representing latent PA and NA factors with acceptable fit (CFI = .98, TLI = .97, RMSEA = .07). The model revealed a significant PA × NA interaction (log-*b* = –0.22, SE = 0.10, *p* = .02; Figure 2E). Higher NA progressively attenuated the positive association between PA and cf-mtDNA. At low levels of NA, higher PA was associated with elevated cf-mtDNA. At the mean level of NA, higher PA was associated with higher cf-mtDNA (log-*b* = .30, SE = 0.11, *p* = .008). When NA was high, there was no relationship between PA and cf-mtDNA (*p* > .05). No significant main or interaction effects were observed for cf-nDNA (log-*b* = –0.15, SE = 0.11, *p* = .16). Participant trajectories for cf-mtDNA and cf-nDNA across time by hourly NA and PA levels are shown in Supplemental Figures S3A–B and S5A–B.

The variance explained was considerably lower in Study 2 compared to Study 1. Across the univariate models, marginal R^2^ values (fixed effects) accounted for approximately 1% to 3% of the variance. Allowing random slopes for happiness and stress modestly increased the explained variance from 1.5% to 2.2% and 1.3% to 1.4%, respectively, with the total variance explained remaining low (2.6% and 1.6%). The model with the greatest explanatory power was the one that tested the PA × NA interaction, which increased the fixed-effects R² to 3% and the total variance to 3.6%.

#### 3.2.5 Sensitivity analysis: Cf-nDNA is related to frustration and happiness

In sensitivity analyses using cf-nDNA, 6 of the 8 emotions had no relation to momentary cf-nDNA. Happiness was associated with higher levels of cf-nDNA (log-*b* = 0.21, SE = 0.07, *p* = .003), and frustration was associated with lower levels of cf-nDNA (log-*b* = –0.16, SE = 0.07, *p* = .02; Supplemental Figures S4A–S4B; Supplemental Table S7). These associations were similar after covariate adjustment (Supplemental Table S8). Results from the PA × NA interaction model were similar to those of the cf-mtDNA model (log-*b* = –0.17, SE = 0.07, *p* = .01; Supplemental Figure S4C), suggesting that elevated negative emotions may dampen the release of both nuclear and mitochondrial DNA at this time scale. Participant cf-nDNA trajectories by NA (Supplemental Figure S5A) and PA (Supplemental Figure S5B) are provided in the Supplement.

## 4. Discussion

This study used both within-person and between-group analyses to evaluate the relationship between saliva cf-mtDNA and naturally occurring stressors and emotions in daily life. Overall, we found converging evidence that saliva cf-mtDNA is sensitive to stress, affect, and MDD diagnosis. At both the day and hourly level, naturalistic stressors were associated with reduced saliva cf-mtDNA concentrations. At the day level, this effect was specific to individuals with MDD and to cf-mtDNA, as it was not replicated with cf-nDNA. Preliminary findings suggest the existence of a dose-response relationship between the number or severity of daily stressors and cf-mtDNA levels. Study 2 replicated and extended the protocol of Study 1, revealing stress-induced attenuation of cf-mtDNA at the hourly level, with significant between-person heterogeneity.

Although prior work on cf-mtDNA has primarily examined psychological stress in isolation, the present study extends this literature by situating stress within a broader affective context. Person-centered analyses demonstrated that higher-than-average positive emotions were associated with higher cf-mtDNA levels, while higher-than-average stress and frustration were associated with lower cf-mtDNA levels. As elevated cf-mtDNA levels have been repeatedly associated with poor health outcomes such as worse illness severity in sepsis (de Miranda et al., 2024; Harrington et al., 2019), reduced survival in cancer (Meng et al., 2019; Sayal et al., 2023), and a higher risk of all-cause mortality in population-based cohorts (Kananen et al., 2020), it is unlikely that positive emotions directly increase cf-mtDNA concentrations per se. Rather, negative psychosocial experiences may limit the normal fluctuations of cf-mtDNA. The emerging picture is one in which daily stressors restrict the range over which saliva cf-mtDNA can fluctuate on short time scales. PA (in the absence of NA) may not increase cf-mtDNA, but permit more flexibility, which may be interpreted as an indication of adaptive capacity.

It should be noted that the direction of the associations observed in the present study diverged from those found in the laboratory (Limberg et al., 2025; Trumpff et al., 2019). This is not entirely unexpected, as real-world stressors are considerably different from those administered in controlled settings (Limberg et al., 2025; Trumpff et al., 2019). Everyday stressors often co-occur, persist for extended periods, and lack clear resolution, decreasing the likelihood of observing discrete peaks in cf-mtDNA. More studies using ambulatory designs are needed, as the stressors that contribute to disease unfold in the diffuse, ongoing conditions of daily life, rather than in brief, well-defined laboratory paradigms. Moreover, recent evidence indicates that saliva cf-mtDNA levels rise rapidly (within 10 minutes) following acute psychosocial stress (Trumpff et al., 2025). Therefore, the probability of obtaining saliva within that narrow post-stressor window with a standardized sampling schedule is low. If daily-life stress produces a similarly brief cf-mtDNA spike followed by a longer trough, then standardized hourly schedules will under-sample peaks and over-sample recovery. This offers a parsimonious explanation for the lower cf-mtDNA concentrations observed on stressor days, motivating event-contingent sampling to map peak-to-nadir trajectories in daily life.

In Study 1, MDD diagnosis alone was not associated with cf-mtDNA, and group differences emerged only in the context of stress exposure. Depressed individuals tend to experience stronger and more persistent emotional responses to everyday challenges, along with slower recovery and greater physiological reactivity (Peeters et al., 2003), which may impact their cf-mtDNA response. A stress-contingent dynamic aligns with diathesis-stress models in which latent vulnerabilities manifest under environmental challenge (Monroe & Simons, 1991). Similar results have been reported in studies of individuals with early-life adversity, in which group-by-gene expression differences only emerge under acute stress (Etzel et al., 2024). Rather than a stable biomarker used to compare groups, cf-mtDNA may be best conceptualized as a dynamic marker of stress-related vulnerability or resilience, more in line with traditional biomarkers of stress, like cortisol, where reactivity to stress, depending on the condition, is increased or decreased (Morris et al., 2014). Cf-mtDNA may also be more similar to heart rate variability, in which reduced variability is associated with poor health and increased variability is associated with better health. This reframing helps explain mixed cross-sectional findings and motivates future work to probe potential mitochondrial stress-recovery phenotypes in psychiatric populations (Melamud et al., 2023; Park et al., 2022).

In Study 2, where saliva was sampled up to 40 times per participant across two days, models incorporating random slopes revealed that the direction and magnitude of the effects were not universal. A minority of participants showed lower cf-mtDNA concentrations with higher levels of happiness, while others showed higher cf-mtDNA with increased stress. These between-person differences may reflect methodological factors. It is possible that these individuals are fundamentally different from their peers in some way. Prior work suggests that situational physiological states and individual differences in blood pressure, affect, and other parameters explain some of the variance in blood cf-mtDNA reactivity (Trumpff et al., 2019), a relationship that has not yet been studied in saliva. Alternatively, participants with positive slopes may have been sampled closer to a discrete stress exposure, which would increase the likelihood of capturing the post-stress influx of cf-mtDNA. To disentangle timing artifacts from true individual differences, future studies should employ designs that trigger saliva collection immediately after stress exposure and estimate person-specific slopes using mixed-effects models, an approach well established in EMA research for capturing rapid, within-person processes in daily life (Dawood et al., 2020).

A key takeaway from this study is that the physiological effects of stress and emotions should not be studied in isolation, and univariate models can oversimplify complex cf-mtDNA dynamics. Examination of Figure 2E demonstrates that the PA-cf-mtDNA relationship was contingent upon the level of NA. The danger of testing the effect of negative emotions on cf-mtDNA without considering the broader affective context is the risk of an incomplete (or even inaccurate) conclusion that particular emotions work singularly on cf-mtDNA. Humans are capable of feeling more than one emotion simultaneously, and it is our duty as researchers not to oversimplify that inner complexity. As this study highlights, there may be nuanced effects that calibrate mitochondrial signaling in nonlinear ways.

Compared to Study 1, where fixed effects explained up to 30.8% of the total variance of cf-mtDNA, the explanatory power of momentary affect for cf-mtDNA in Study 2 was smaller. This likely reflects both the finer temporal resolution and greater volatility of within-day fluctuations, which reduce the proportion of variance attributable to single predictors when averaged at the group level. Despite the relatively small proportion of average variance explained by the fixed effects in Study 2, these values are consistent with those found in other intensive longitudinal EMA studies (McNeish et al., 2021). Hour-to-hour assessments introduce substantial biological and contextual noise as unmeasured variables (e.g, sleep, diet, illness, or physical activity), which likely influence cf-mtDNA levels. In this context, even small percentages of explained variance can reflect a meaningful biological signal. By contrast, Study 1 yielded larger fixed-effects R^2^ values, likely because day-level averaging reduced noise. Despite the explanatory power of momentary affect being smaller at the hourly level, these effects are not necessarily inconsequential. Instead, they highlight the sensitivity of cf-mtDNA to psychological states even against a backdrop of high within-person volatility. While these findings indicate that cf-mtDNA shows detectable associations with psychological stress at the day level, denser sampling is likely necessary to fully characterize the underlying temporal dynamics.

To contextualize these patterns, we compared cf-mtDNA with cf-nDNA. Cf-mtDNA consistently demonstrated greater sensitivity to psychological states than cf-nDNA, corroborating findings from previous studies (Behnke et al., 2025). In Study 1, cf-nDNA showed no relationship with stress exposure or the number or severity of stressors. This divergence supports the idea that a portion of cf-mtDNA is released through an active mechanism independent of cell death. Although cf-mtDNA and cf-nDNA were moderately correlated in this sample, only cf-mtDNA was associated with stress, suggesting differential biological sensitivity despite shared variance. In Study 2, cf-mtDNA again demonstrated broader emotional reactivity, correlating with six of the eight momentary affective states, while cf-nDNA was associated with only two affective states. Notably, cf-mtDNA and cf-nDNA were strongly correlated in Study 2, yet their associations with emotional states diverged, highlighting how cf-mtDNA captured unique variance.

While this remains to be confirmed by additional studies, the observed decrease in cf-mtDNA during stress may reflect an adaptive suppression of mitochondrial signaling. From a systems perspective, this could serve as a stabilizing strategy that helps conserve energy, dampen excessive signaling cascades, and maintain homeostasis when an organism is threatened (Bobba-Alves, Juster & Picard, 2022). Resilience often involves the selective silencing or attenuation of biological reactivity when sustained activation would be metabolically costly or maladaptive. This energy conservation model aligns well with theories such as allostatic load and our emerging understanding of the aging process (Shaulson, Cohen & Picard, 2024). This interpretation remains speculative and requires direct testing to confirm.

### 4.1 Limitations

By conventional standards, both cohorts were modest in size; however, each study employed intensive repeated-measures designs, which increased the precision and reliability of within-person estimates. It should be noted that this design does not eliminate limitations related to between-subject sample size or generalizability. With the feasibility of intensive saliva cf-mtDNA established, these associations should be tested in larger and more diverse cohorts.

Although Study 2 utilized frequent measurement of saliva cf-mtDNA, the temporal resolution may still have been too coarse to capture these mechanisms as they occur in daily life. Data regarding the precise timing of stressors relative to saliva sampling was unavailable, which may explain the absence of short-lag cf-mtDNA spikes. To better understand the true mitochondrial dynamics that follow naturalistic stressors, future research should integrate event-contingent saliva sampling protocols to reveal for whom and under what circumstances cf-mtDNA is released or suppressed. Such approaches will likely enhance our understanding of the psychological and contextual factors shaping mitochondrial signaling and clarify its role in stress reactivity and resilience.

Finally, Study 2 presumes that the average level of each emotion reported across a two-day period reflects one’s true affective baseline. The person-centered values were derived using data from only two days, and variation from these values indicated meaningful fluctuations in affect. However, a baseline derived from two days of data may not accurately capture one’s normative state, underscoring the need for longer sampling periods. Despite their limitations, differences in sampling intensity, and cohort characteristics., both studies showed effects in the same direction.

### 4.2 Conclusion

Here, we show that cf-mtDNA measured in daily life does not exhibit a uniform or directionally consistent association with psychological states. Across two intensive longitudinal studies, cf-mtDNA levels varied as a function of temporal context, affective state, and individual differences, rather than showing a simple stress-reactive increase. These findings do not contradict prior laboratory work. Rather, they suggest that real-world mitochondrial signaling unfolds on time scales and under conditions that are not captured by coarse or fixed sampling schedules. This study contributes to a developing area of research demonstrating cf-mtDNA as a dynamic, context-dependent biomarker that is highly sensitive to timing and measurement design.

## Data Availability

All data produced in the present study are available upon reasonable request to the authors

## CRediT

Lauren Petri: Conceptualization, Formal Analysis, Writing - Original Draft

Sun Ah Lee: Data Curation, Writing - review and editing

David Shire: Visualization, Investigation, Writing - review and editing

Samantha Leonard: Investigation, Writing - review and editing

Alexander Behnke: Writing - review and editing

Jody Greaney: Resources, Writing - review and editing

Lacy Alexander: Resources, Writing - review and editing

David Almeida: Conceptualization, Supervision, Writing - review and editing

Martin Picard: Supervision, Resources, Writing - review and editing

Caroline Trumpff: Conceptualization, Formal Analysis, Writing - Original Draft, Supervision, Project Administration.

