## Supplemental Table S1 for "Saliva cell-free mitochondrial DNA (cf-mtDNA) as a dynamic biomarker of stress and emotion in daily life: Evidence from two independent repeated-measures studies"

| **Table S1** |  |  |
| --- | --- | --- |
| *Study 1: Sensitivity analysis of day-level stress predicting cf-mtDNA in sequential GLMMs (statistical outlier included)* | | |
| Predictor | Model 1 | Model 2 |
| Fixed effects |  |  |
| Intercept | 8.66 (0.29)*** | 4.88 (1.55)** |
| Stress | –0.67 (0.12)*** | –0.66 (0.12)*** |
| Group (MDD) | –0.52 (0.44) | –0.55 (0.37) |
| Stress × Group | –0.35 (0.21) | –0.36 (0.21)*** |
| Age | — | 0.12 (0.05)* |
| BMI | — | 0.04 (0.04) |
| Sex (Male) | — | 0.55 (0.37) |
| Random Effects |  |  |
| Intercept variance | 0.77 (0.88) | 0.53 (0.73) |
| Model Fit Indices |  |  |
| AIC | 4,708 | 4,708 |
| BIC | 4,730 | 4,740 |
| *Note.* Statistical outlier included in generalized linear mixed-effects models (GLMMs) with a gamma error distribution, a log link function, and random subject intercepts using stress exposure to predict day-level cell-free mitochondrial DNA (cf-mtDNA). Model 1 tests the stress × group interaction; Model 2 adjusts for covariates age, BMI, and sex. Coefficients are shown as Estimate (SE). Model fit statistics (AIC & BIC) are reported for model comparison. * *p* < .05*, ** p < .01, *** p < .001*. | | |
