## Supplemental Table S2 for "Saliva cell-free mitochondrial DNA (cf-mtDNA) as a dynamic biomarker of stress and emotion in daily life: Evidence from two independent repeated-measures studies"

| **Table S2** | | |
| --- | --- | --- |
| *Study 1: Day-level stress exposure predicting cf-nDNA* | | |
| Predictor | Model 1 | Model 2 |
| Fixed effects |  |  |
| Intercept | 6.05 (0.36)*** | 2.74 (1.66) |
| Stress | –0.08 (0.32) | –0.04 (0.32) |
| Group (MDD) | –0.33 (0.53) | –0.46 (0.47) |
| Stress × Group | –0.56 (0.51) | –0.44 (0.49) |
| Age | — | 0.14 (0.05)** |
| BMI | — | 0.006 (0.04) |
| Sex (Male) | — | 0.34 (0.40) |
| Random Effects |  |  |
| Intercept variance | 0.73 (0.86) | 0.46 (0.68) |
| Model Fit Indices |  |  |
| AIC | 913 | 913 |
| BIC | 926 | 932 |
| *Note*. Replication of Model 1 using cf-nDNA. Generalized linear mixed-effects models (GLMMs) with a gamma error distribution, a log link function, and random subject intercepts using stress exposure to predict day-level cell-free nuclear DNA (cf-nDNA). Model 1 tests the stress × group interaction; Model 2 adjusts for covariates age, BMI, and sex. Coefficients are shown as Estimate (SE). Model fit statistics (AIC & BIC) are reported for model comparison. * *p* < .05*, ** p < .01, *** p < .001*. | | |
