## Supplemental Table S3 for "Saliva cell-free mitochondrial DNA (cf-mtDNA) as a dynamic biomarker of stress and emotion in daily life: Evidence from two independent repeated-measures studies"

| **Table S3** |  |  |
| --- | --- | --- |
| *Study 1: Dose-response effect of stress, number and severity of stressors predicting cf-mtDNA* | | |
| Predictor | Model 1 Number | Model 2 Severity |
| Fixed effects |  |  |
| Intercept | 8.23 (0.22)*** | 8.26 (0.23)*** |
| Stress | –0.31 (0.12)* | –0.28 (0.10)* |
| Random Effects |  |  |
| Intercept variance ID | 0.56 (0.75) | 0.62 (0.79) |
| Model Fit Indices |  |  |
| AIC | 1,184 | 1,182 |
| BIC | 1,193 | 1,191 |
| *Note.* Results from generalized linear mixed-effects models (GLMMs) with a gamma error distribution, a log link function, and random subject intercepts testing stress number and severity to predict day-level cell-free mitochondrial DNA (cf-mtDNA). Model 1 evaluates cf-mtDNA as a function of stressor accumulation; Model 2 evaluates cf-mtDNA as a function of stressor severity. Coefficients are shown as Estimate (SE). Model fit statistics (AIC & BIC) are reported for comparison. * *p* < .05*, ** p < .01, *** p < .001*. | | |
