## Supplemental Table S4 for "Saliva cell-free mitochondrial DNA (cf-mtDNA) as a dynamic biomarker of stress and emotion in daily life: Evidence from two independent repeated-measures studies"

| **Table S4**  *Study 2: Person-centered psychological states predicting cf-mtDNA in GLMMs with random intercepts and covariates* | | | | | | | | |
| --- | --- | --- | --- | --- | --- | --- | --- | --- |
|  | Anxious | Calm | Control | Energetic | Fatigued | Frustrated | Happy | Stress |
| Fixed Effects | |  |  |  |  |  |  |  |
| Intercept | 8.93 (1.64)*** | 8.92 (1.63)*** | 8.95 (1.62)*** | 8.95 (1.63)*** | 8.94 (1.64)*** | 8.93 (1.64)*** | 8.81 (1.61)*** | 8.96 (1.68)*** |
| Variable | –0.02 (0.06) | 0.15 (0.05)** | 0.15 (0.06)** | 0.16 (0.05)** | –0.01 (0.05) | –0.14 (0.06)* | 0.26 (0.06)*** | –0.16 (0.06)* |
| Age | 0.01 (0.02) | 0.01 (0.02) | 0.01 (0.02) | 0.01 (0.02) | 0.01 (0.02) | 0.01 (0.02) | 0.01 (0.02) | 0.01 (0.02) |
| Sex | 0.77 (0.38)* | 0.77 (0.37)* | 0.76 (0.37)* | 0.77 (0.37)* | 0.77 (0.38)* | 0.77 (0.38)* | 0.75 (0.37)* | 0.79 (0.39)* |
| BMI | 0.03 (0.06) | 0.03 (0.06) | 0.03 (0.06) | 0.03 (0.06) | 0.03 (0.06) | 0.03 (0.06) | 0.03 (0.06) | 0.02 (0.06) |
| Random Effects | |  |  |  |  |  |  |  |
| Variance | 0.80 | 0.79 | 0.78 | 0.79 | 0.80 | 0.80 | 0.77 | 0.84 |
| Model Fit Indices | |  |  |  |  |  |  |  |
| AIC | 17,681 | 17,673 | 17,674 | 17,670 | 17,680 | 17,676 | 17,661 | 16,695 |
| BIC | 17,713 | 17,705 | 17,707 | 17,703 | 17,713 | 17,708 | 17,693 | 16,727 |
| *Note.* Gamma generalized linear mixed-effects models (GLMMs) with random intercepts test the effect of person-centered momentary psychological states on cell-free mitochondrial DNA (cf-mtDNA), excluding the identified statistical outlier. Each model includes a single psychological state (e.g., calm, happy, stressed) as a fixed effect and adjusts for covariates (age, sex, BMI). Findings are consistent with the primary cf-mtDNA models. Model-fit statistics (AIC & BIC) are reported for model comparison. Coefficients are shown as Estimate (SE). **p < .05, **p < .01, ***p < .001.* | | | | | | | | |
