## Supplemental Table S5 for "Saliva cell-free mitochondrial DNA (cf-mtDNA) as a dynamic biomarker of stress and emotion in daily life: Evidence from two independent repeated-measures studies"

| **Table S5** | | | |  |  |  |  |  |
| --- | --- | --- | --- | --- | --- | --- | --- | --- |
| *Study 2: Person-centered psychological states predicting cf-mtDNA in GLMMs with random intercepts (statistical outlier included)* | | | | | | | | |
|  | Anxious | Calm | Control | Energetic | Fatigued | Frustrated | Happy | Stressed |
| Fixed Effects | |  |  |  |  |  |  |  |
| Intercept | 10.32 (0.21)*** | 10.32 (0.21)*** | 10.32 (0.21)*** | 10.31 (0.21)*** | 10.32 (0.21)*** | 10.32 (0.21)*** | 10.31 (0.20)*** | 10.32 (0.21)*** |
| Variable | –0.02 (0.07) | 0.16 (0.005)** | 0.16 (0.06)** | 0.16 (0.05)** | –0.01 (0.05) | –0.14 (0.06)* | 0.26 (0.06)*** | –0.16 (0.06)* |
| Random Effects | | | |  |  |  |  |  |
| Variance | 1.04 (1.02) | 1.03 (1.01) | 1.02 (1.01) | 1.02 (1.01) | 1.04 (1.02) | 1.04 (1.02) | 1.00 (1.00) | 1.09 (1.04) |
| Model Fit Indices | |  |  |  |  |  |  |  |
| AIC | 18,383 | 18,375 | 18,375 | 18,373 | 18,383 | 18,378 | 18,363 | 17,355 |
| BIC | 18,402 | 18,394 | 18,394 | 18,392 | 18,402 | 18,397 | 18,382 | 17,373 |
| *Note.* Gamma generalized linear mixed-effects models (GLMMs) with random intercepts test the effect of person-centered momentary psychological states on cell-free mitochondrial DNA (cf-mtDNA), including the identified statistical outlier. Person-centered scores are computed by centering each state relative to the individual’s mean. Each model includes a single psychological variable as a fixed effect. Model-fit statistics (AIC & BIC) are reported for model comparison. Coefficients are shown as Estimate (SE). **p < .05, **p < .01, ***p < .001.* | | | | | | | | |
