## Supplemental Table S6 for "Saliva cell-free mitochondrial DNA (cf-mtDNA) as a dynamic biomarker of stress and emotion in daily life: Evidence from two independent repeated-measures studies"

| **Table S6** | |  |  |  |  |  |  |  |
| --- | --- | --- | --- | --- | --- | --- | --- | --- |
| *Study 2: Person-centered psychological states predicting cf-mtDNA in GLMMs with random intercepts and random slopes* | | | | | | | | |
|  | Anxious | Calm | Control | Energetic | Fatigued | Frustrated | Happy | Stressed |
| Fixed Effects | |  |  |  |  |  |  |  |
| Intercept | 10.27(0.21)*** | 10.23 (0.20)*** | 10.28 (0.20)*** | 10.26 (0.20)*** | 10.29 (0.20)*** | 10.28 (0.20)*** | 10.23 (0.20)*** | 10.25 (0.21)*** |
| Variable | –0.12 (0.13) | 0.19 (0.13) | 0.13 (0.08) | 0.16 (0.09) | –0.02 (0.07) | –0.19 (0.12) | 0.31 (0.12)** | –0.28 (0.12)* |
| Random Effects | | | |  |  |  |  |  |
| Variance_i_ | 1.01 (1.01) | 0.99 (0.99) | 0.98 (0.99) | 0.97 (0.98) | 0.99 (1.00) | 1.01 (1.00) | 0.96 (0.98) | 1.09 (1.05) |
| Variance_s_ | 0.20 (0.44) | 0.30 (0.55) | 0.06 (0.25) | 0.13 (0.36) | 0.04 (0.20) | 0.14 (0.37) | 0.24 (0.49) | 0.19 (0.43) |
| Correlation | –0.30 | 0.14 | 0.48 | 0.10 | –0.03 | –0.04 | 0.19 | 0.08 |
| Model Fit Indices | |  |  |  |  |  |  |  |
| AIC | 18,303 | 18,271 | 18,305 | 18,297 | 18,320 | 18,309 | 18,268 | 17,264 |
| BIC | 18,331 | 18,299 | 18,333 | 18,325 | 18,348 | 18,337 | 18,296 | 17,292 |
| *Note.* Gamma generalized linear mixed-effects models (GLMMs) with random intercepts and slopes test the effect of person-centered momentary psychological states on cell-free mitochondrial DNA (cf-mtDNA). Person-centered scores are computed by centering each state on the individual’s mean. Variance estimates for intercepts and slopes, along with their correlations, reflect individual differences in baseline cf-mtDNA levels and in the strength of associations with psychological states. Model-fit statistics (AIC & BIC) are reported for comparison. Coefficients are shown as Estimate (SE). **p < .05, ***p < .01, **p < .001.* | | | | | | | | |
