## Supplemental Table S7 for "Saliva cell-free mitochondrial DNA (cf-mtDNA) as a dynamic biomarker of stress and emotion in daily life: Evidence from two independent repeated-measures studies"

| **Table S7** | |  |  |  |  |  |  |  |
| --- | --- | --- | --- | --- | --- | --- | --- | --- |
| *Study 2: Person-centered psychological states predicting cf-nDNA in GLMMs with random intercepts* | | | | | | | | |
|  | Anxious | Calm | Control | Energetic | Fatigued | Frustrated | Happy | Stressed |
| Fixed Effects | | | | | | | | |
| Intercept | 6.63 (0.22)*** | 6.62 (0.22)*** | 6.62 (0.22)*** | 6.62 (.22)*** | 6.62 (0.22)*** | 6.62 (0.22)*** | 6.61 (0.22)*** | 6.63 (0.22)*** |
| Variable | –0.01 (0.08) | 0.11 (0.06) | 0.12 (0.07) | .07 (.06) | –0.09 (0.06) | –0.16 (0.07)* | 0.21 (0.07)** | –0.11 (0.07) |
| Random Effects | | | |  |  |  |  |  |
| Variance | 1.16 (1.08) | 1.15 (1.07) | 1.14 (1.07) | 1.16 (1.07) | 1.16 (1.08) | 1.17 (1.08) | 1.15 (1.07) | 1.17 (1.08) |
| Model Fit Indices | | |  |  |  |  |  |  |
| AIC | 11,329 | 11,326 | 11,325 | 11,351 | 11,326 | 11,324 | 11,320 | 10,661 |
| BIC | 11,347 | 11,344 | 11,344 | 11,370 | 11,345 | 11,342 | 11,338 | 10,679 |
| *Note.* Gamma generalized linear mixed-effects models (GLMMs) with random intercepts test the effect of person-centered momentary psychological states on cell-free nuclear DNA (cf-nDNA), excluding the identified statistical outlier. Person-centered scores are computed by centering each state on the individual’s mean. Frustration and happiness significantly predict cf-nDNA, whereas other states are not associated. Model-fit statistics (AIC & BIC) are reported for comparison. Coefficients are shown as Estimate (SE). **p < .05, **p < .01, ***p < .001.* | | | | | | | | |
