## Supplemental Table S8 for "Saliva cell-free mitochondrial DNA (cf-mtDNA) as a dynamic biomarker of stress and emotion in daily life: Evidence from two independent repeated-measures studies"

| **Table S8**  *Study 2: Person-centered psychological states predicting cf-nDNA in GLMMs with random intercepts models and covariates* | | | | | | | | |
| --- | --- | --- | --- | --- | --- | --- | --- | --- |
|  | Anxious | Calm | Control | Energetic | Fatigued | Frustrated | Happy | Stress |
| Fixed Effects | |  |  |  |  |  |  |  |
| Intercept | 2.84 (1.78) | 2.84 (1.77) | 2.85 (1.76) | 2.85 (1.77) | 2.86 (1.78) | 2.84 (1.78) | 2.72 (1.76) | 2.85 (1.78) |
| Variable | 0.01 (0.08) | 0.11 (0.06) | 0.12 (0.07) | 0.08 (0.06) | 0.08 (0.06) | –0.16 (0.07)* | 0.22 (0.07)** | –0.12 (0.07) |
| Age | 0.03 (0.03) | 0.03 (0.03) | 0.03 (0.03) | 0.03 (0.03) | 0.03 (0.03) | 0.03 (0.03) | 0.03 (0.03) | 0.03 (0.03) |
| Sex | 0.62 (0.41) | 0.61 (0.41) | 0.60 (0.41) | 0.61 (0.41) | 0.61 (0.41) | 0.62 (0.41) | 0.58 (0.41) | 0.64 (0.41) |
| BMI | 0.11 (0.07) | 0.11 (0.07) | 0.11 (0.07) | 0.11 (0.07) | 0.11 (0.07) | 0.11 (0.07) | 0.12 (0.07) | 0.11 (0.07) |
| Random Effects | |  |  |  |  |  |  |  |
| Variance | 0.93 | 0.93 | 0.92 | 0.93 | 0.94 | 0.94 | 0.92 | 0.94 |
| Model Fit Indices | |  |  |  |  |  |  |  |
| AIC | 11,079 | 11,076 | 11,076 | 11,077 | 11,077 | 11,074 | 11,070 | 10,438 |
| BIC | 11,111 | 11,108 | 11,108 | 11,109 | 11,109 | 11,106 | 11,102 | 10,470 |
| *Note.* Gamma generalized linear mixed-effects models (GLMMs) with random intercepts test the effect of person-centered momentary psychological states on cell-free nuclear DNA (cf-nDNA), excluding the identified statistical outlier. Each model includes a single psychological variable as a fixed effect and covariates (age, sex, BMI). Person-centered scores are computed by centering each state on the individual’s mean. Model-fit statistics (AIC & BIC) are reported for comparison. Coefficients are shown as Estimate (SE). **p < .05, **p < .01, ***p < .001.* | | | | | | | | |
