## Supplemental Figure for "Saliva cell-free mitochondrial DNA (cf-mtDNA) as a dynamic biomarker of stress and emotion in daily life: Evidence from two independent repeated-measures studies"

**Figure S1**

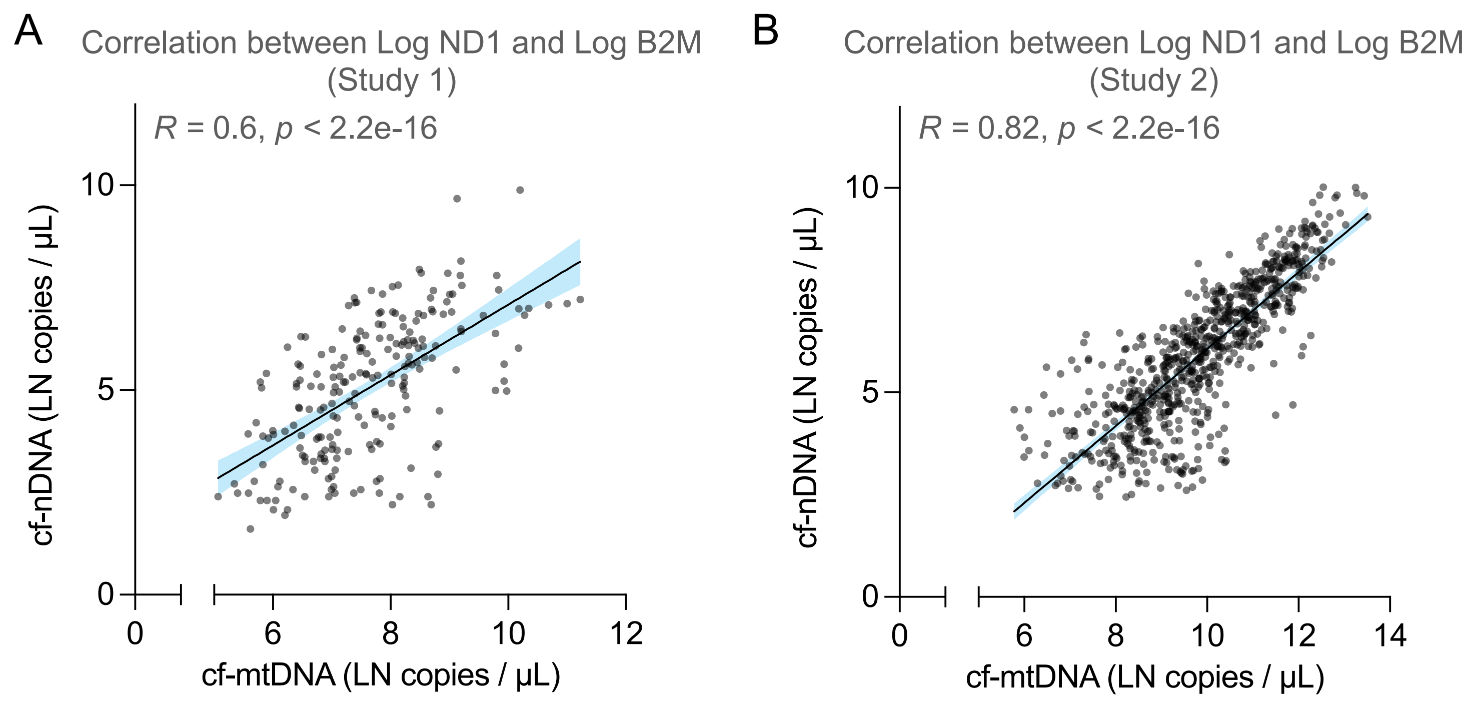


Supplemental Figure 1. The relationship between the log cell-free mitochondrial DNA (cf-mtDNA) and cell-free nuclear DNA (cf-nDNA) across repeated measurements in both studies. **(A)** For study 1 there was a significant repeated measures correlation (r = .60, df = 199, p < .001). **(B)** In Study 2, results from a repeated measures correlation suggest that cf-mtDNA and cf-nDNA were highly correlated ($r_{rm}$ = 0.81, df = 725, 95% CI [.79, .84], p < .001).

**Figure S2**

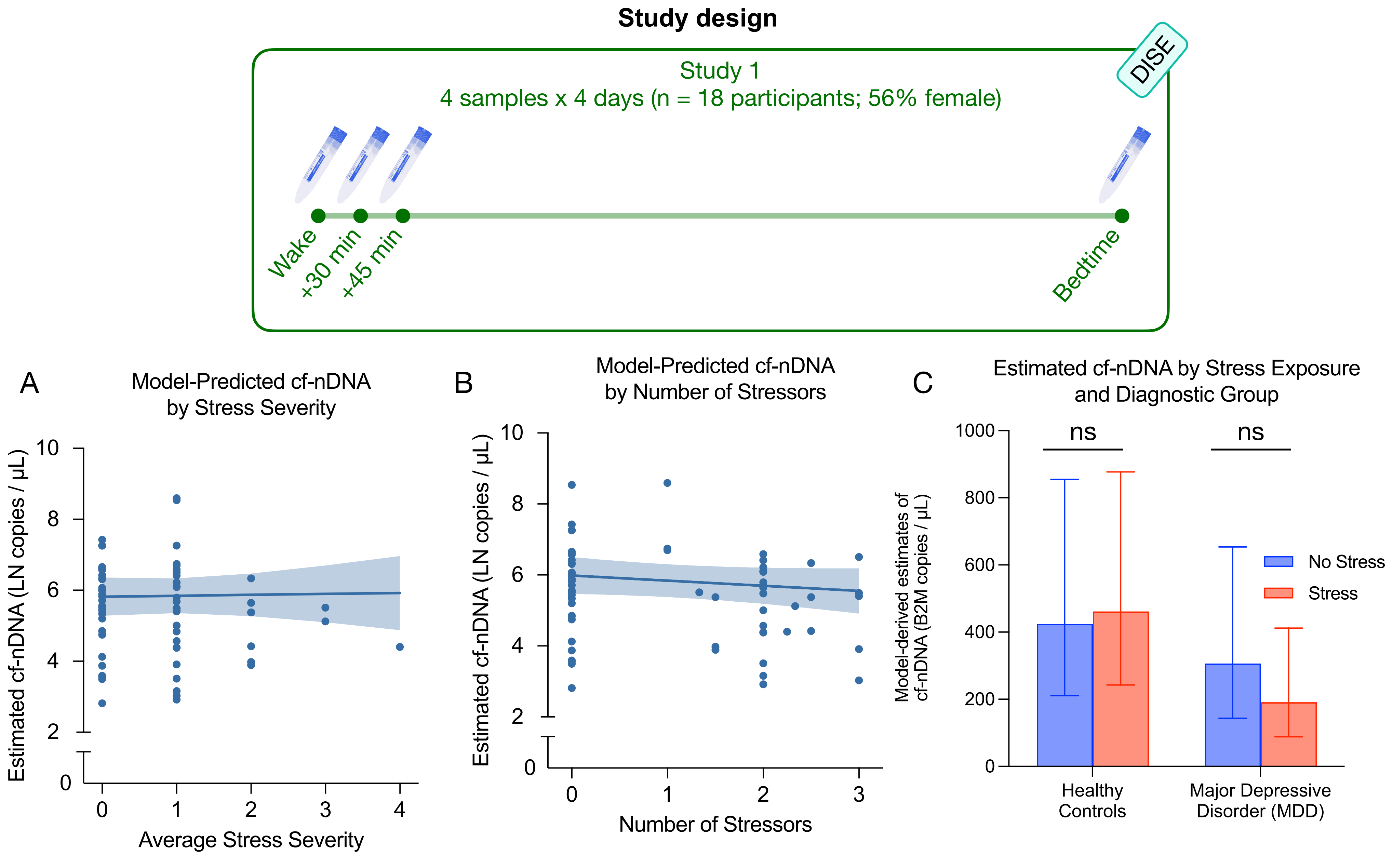


Supplemental Figure 2 Study 1. **(A, B)** Model-predicted values of cf-nDNA as the number of stressors reported and the average stress severity increase. **(C)** The interaction between stressor and non-stressor days for healthy controls and participants with major depressive disorder.

**Figure S3**

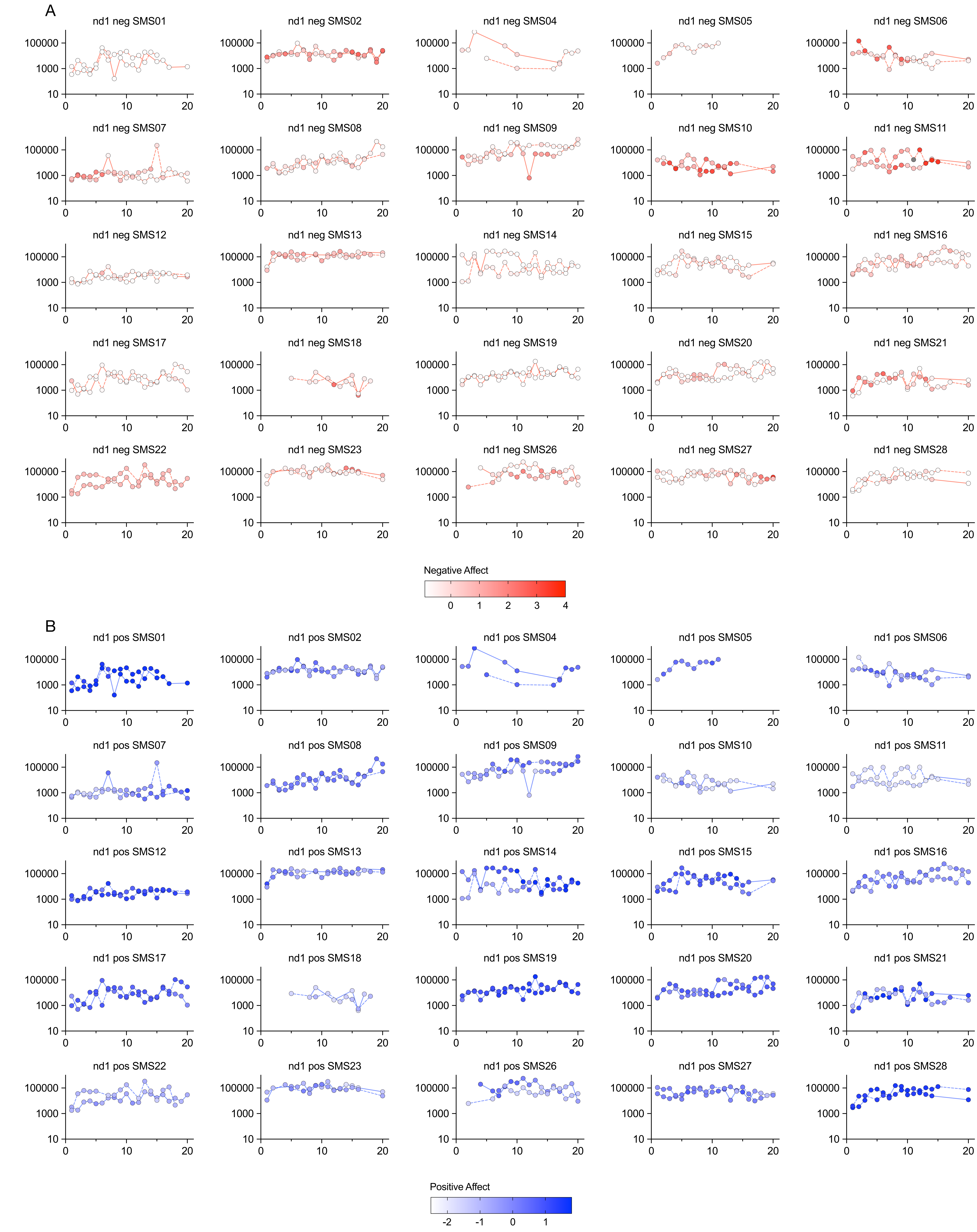


Supplemental Figure 4. **(A)** Cf-mtDNA by Negative Affect Intensity and **(B)** Positive Affect intensity over the course of the day

**Figure S4**

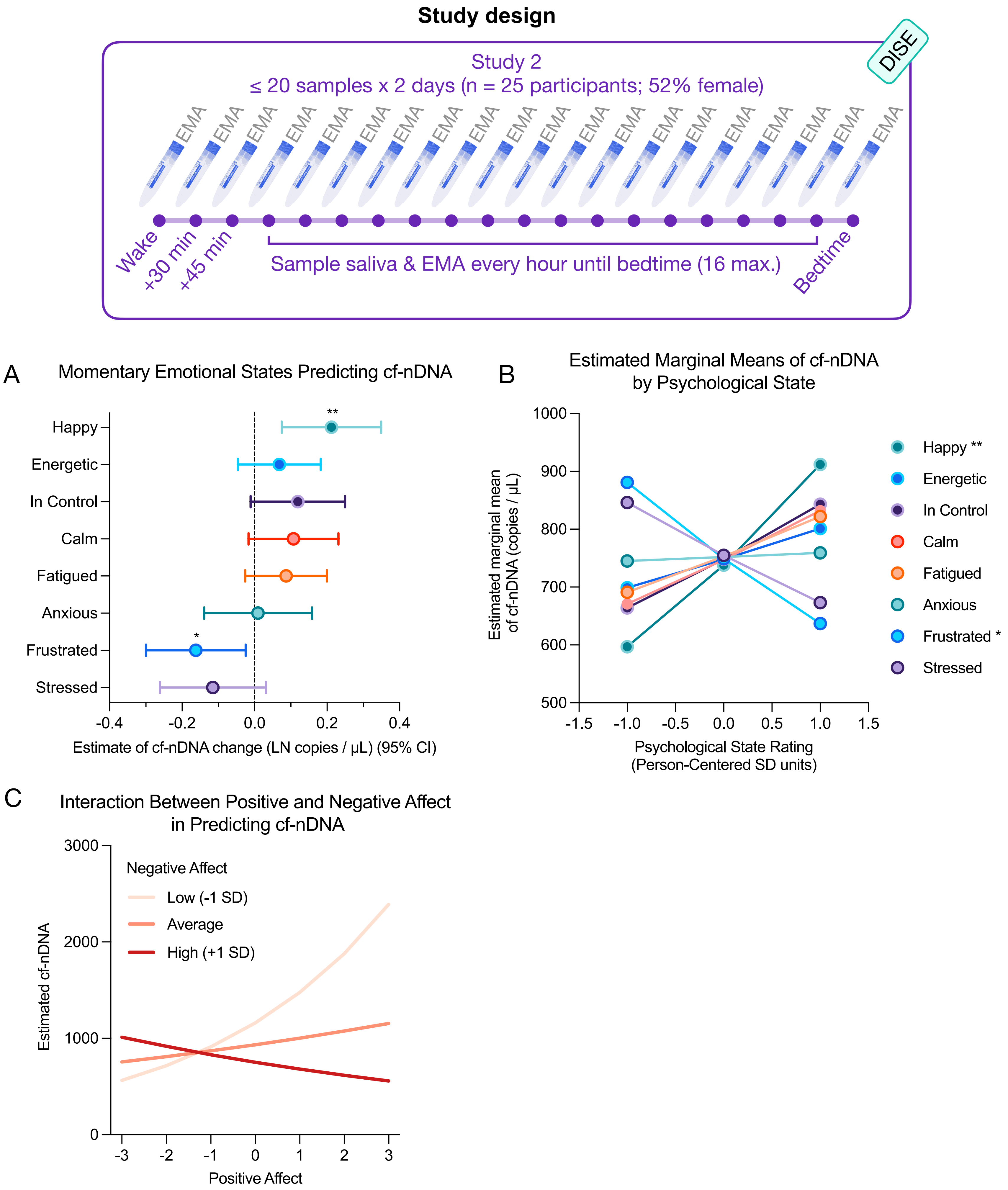


Supplemental Figure 3. **Replication of cell-free mitochondrial DNA (cf-mtDNA) models with cell-free nuclear DNA (cf-nDNA) in Study 2. (A)** Forest plot of estimates from generalized linear mixed-effects models across all 8 participant-centered bivariate models for cf-nDNA. **(B)** Estimated marginal means of cf-nDNA levels by psychological state (+/- 1 SD). **(C)** Interaction between positive and negative affect latent factors on cf-nDNA.

**Figure S5**

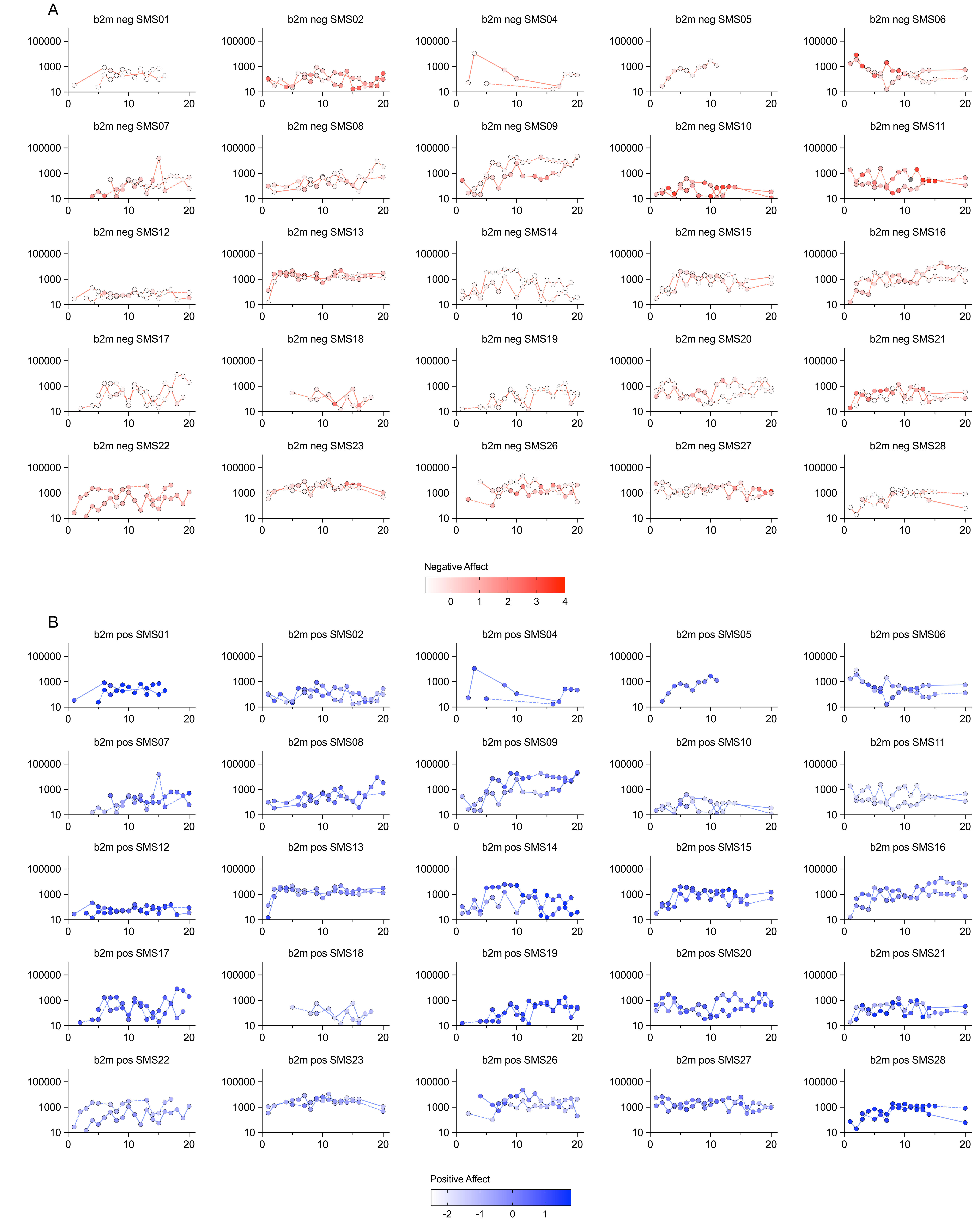


Supplemental Figure 4. **(A)** Cf-nDNA by Negative Affect Intensity and **(B)** Positive Affect intensity over the course of the day
